# Integrating Bioinformatics and Machine Learning to Identify Mitochondria-Related Biomarkers and Their Association with Immune Infiltration in BK polyomavirus-associated nephropathy

**DOI:** 10.1101/2025.07.07.25331061

**Authors:** Changhai Xu, Xueying Wang, Haibo Wu, Wei Li, Fei Lin, Na Lin, Shiyin Shen, Shubin Pan, Tong Chen, Donghui Zhang, Long He, Yan Cui

## Abstract

BK polyomavirus-associated nephropathy (BKPyVAN) is a serious complication of kidney transplantation. Numerous kidney diseases such as BKPyVAN have been shown to cause mitochondrial dysfunction. This study aims to identify key mitochondria-related genes in BKPyVAN. We merged two datasets, GSE72925 and GSE47199, to form a training set after batch-effect removal. Hub mitochondria-related genes in BKPyVAN were identified using bioinformatics tools. The functional information of the hub genes was analyzed using gene set enrichment analysis (GSEA). A mouse model of polyomavirus (MPyV) infection was established to verify the expression levels of B-cell lymphoma 2-related protein A1 (BCL2A1), Caspase-3 (CASP3), and threonine synthase like 1 (THNSL1) in BKPyVAN. We identified nine mitochondria-related genes that were differentially expressed between BKPyVAN and stable graft samples and correlated with BKPyVAN onset. Among these, three genes (*THNSL1*, *BCL2A1*, and *CASP3*) were identified as robust mitochondria-related genes in BKPyVAN. *THNSL1* and *BCL2A1* were upregulated and *CASP3* was downregulated in BKPyVAN samples. Moreover, *THNSL1* and *BCL2A1* were upregulated and *CASP3* was downregulated in MPyV kidney tissue samples. These genes showed a significant diagnostic value for BKPyVAN. GSEA revealed the potential involvement of these genes in the immune pathways. Additionally, we found correlations between these genes and immune cell infiltration in BKPyVAN patients. Our study identified three robust biomarkers (*THNSL1*, *BCL2A1* and *CASP3*) for BKPyVAN that might be potential targets for diagnosis and treatment. These biomarkers may be involved in immune pathways and show a significant correlation with immune cell infiltration, suggesting their critical role in BKPyVAN pathogenesis.

## 1 Introduction

Kidney transplantation is a lifesaving procedure that serves as the sole alternative to lifelong dialysis for patients in the final stage of kidney dysfunction, specifically uremia or end-stage renal disease (ESRD) (1). Recipients of kidney transplants are required to take potent immunosuppressive medications to prevent rejection of the transplanted organ (2). However, immunosuppressive therapy can lead to the reactivation of BK polyomavirus (BKPyV) in these individuals, resulting in BK polyomavirus-associated nephropathy (BKPyVAN) (3). BKPyVAN occurs in approximately 1–10% of kidney transplant recipients (4) and is associated with graft dysfunction in over 90% of cases, as well as graft loss in more than 50% of affected individuals (3). To date, there are no effective antiviral prevention strategies or validated treatment options for BKPyV infection. Therefore, there is an urgent need to discover accurate and reliable diagnostic biomarkers for BKPyVAN.

Mitochondria are indispensable organelles that maintain cellular energy metabolism and signal organelles to maintain their biological functions. Mitochondrial dysfunction has been observed in many kidney diseases, including BKPyVAN. Gao et al. observed a significant reduction in both the concentration and composition of urinary cell-free mitochondrial DNA (cfmtDNA) in patients with BKPyVAN and a decrease in the volume of mitochondria within the renal tubular cells of these patients (5). A recent study utilizing single-cell RNA sequencing revealed that oxidative phosphorylation and mitochondrial dysfunction are the top canonical pathways in BKPyVAN patients (6). Manzetti et al. demonstrated that the small accessory agarose protein expressed by BKPyV facilitates BKPyV replication by disrupting mitochondrial membrane potential, increasing autophagic flux, enhancing the availability of biosynthetic building blocks, and fragmentation of the mitochondrial network surrounding the nucleus (7). Furthermore, mitochondrial fragmentation and p62/SQSTM1-mediated autophagy have been observed in BKPyVAN allograft biopsies (7). Therefore, the development of specific mitochondrial factors such as BKPyVAN biomarkers may be of great significance for the study of mitochondrial drug targets for the development of anti-BKPyVAN drugs.

In this study, we identified three mitochondria-related genes as biomarkers for BKPyVAN by integrating bioinformatic approaches with multiple machine learning algorithms. We also evaluated the potential mechanisms by which these three genes may influence BKPyVAN. Furthermore, we analyzed the relationship between hub gene expression and immune cell infiltration in patients with BKPyVAN. This study provides a new scientific basis for the prevention and treatment of BKPyVAN by elucidating the relationship between mitochondrial function and BKPyVAN expression.

## 2 Materials and methods

### 2.1 Data collection

Four datasets related to BKPyVAN and stable grafts (STA), including GSE72925 (BKPyVAN=10, STA=73), GSE47199 (BKPyVAN =3, STA=11), GSE120495 (BKPyVAN =5, STA=5) and GSE75693 (BKPyVAN =15, STA=30) were obtained from the Gene Expression Omnibus (GEO, https://www.ncbi.nlm.nih.gov/geo/) database. Then, the GSE72925 and GSE47199 datasets were merged and used as the training set after removing batch effects via the and “sva” packages in R packages. GSE120495 and GSE75693 were used as external validation datasets.

1136 human mitochondria-related genes were downloaded from the MitoCarta 3.0 database (http://www.broadinstitute.org/mitocarta) (Table S1).

### 2.2 Identification and functional annotation of differentially expressed genes (DEGs)

In the training set, the DEGs between the BKPyVAN and STA groups were identified using “limma” (version 3.56.2) (8) in the R language, based on |Log2FC|>0.58 and FDR<0.05. Subsequently, Gene Ontology (GO: Biological Process (BP), Molecular Function (MF), and Cellular Component (CC)) and Kyoto Encyclopedia of Genes and Genomes (KEGG) enrichment analyses were performed on DEGs using “clusterProfiler” (version 4.8.3) (9) function package in R language. Significantly enriched GO and KEGG pathways were screened using p<0.05.

### 2.3 Weighted gene co-expression network analysis

The analysis utilizing WGCNA was conducted through the “WGCNA” package function (10) in the R programming environment, focusing on gene expression values. Genes were filtered for WGCNA by retaining the top 50% based on the variance analysis. The Pearson correlation coefficient was computed for each gene, allowing for the selection of an appropriate soft threshold β, thus enhancing the network’s adherence to the characteristics of a scale-free network. A one-step approach was implemented to develop a gene network in which the adjacency matrix was transformed into a topological overlap matrix (TOM). Subsequently, hierarchical clustering was employed to create a hierarchical clustering tree representing the genes. To assess the significance of genes in relation to clinical data, gene and module significance were evaluated, and the correlation between modules and traits was thoroughly analyzed to identify crucial gene modules.

### 2.4 Gene set enrichment analysis (GSEA)

In the training dataset, BKPyVAN samples were categorized into groups with high and low expression based on the median levels of each critical gene. GSEA between these two groups was performed using the ClusterProfiler” function package (version 4.8.3) (9) in R language. Significantly enriched pathways were identified at P <0.05.

### 2.5 Identification of key mitochondria-related genes by machine learning

Three machine learning methods, random forest (RF), LASSO, and recursive feature elimination (RFE), were used to screen mitochondria-related genes. The random forest (version 4.6) package in R was applied to build the RF model and select the best feature set. LASSO was performed by “glmnet” package in R language. The “svm” package was used for filter variables using the RFE method. Next, the overlapping genes identified by RF, LASSO, and SVM were considered as key mitochondria-related genes in BKPyV samples.

### 2.6 Immune cell infiltration

CIBERSORT (11) was used to calculate the relative proportions of 22 types of immune cells within the sample. This tool employs a deconvolution algorithm that leverages 547 preset barcode genes to characterize the composition of immune-infiltrating cells based on the gene expression matrix. The total estimated immune cell type proportion in each sample was equal to 1.

### 2.7 Build a ceRNA network

The miRDB (https://mirdb.org/), miRNet (https://www.mirnet.ca/miRNet/home.xhtml), and miRWalk Home-miRWalk (uni-heidelberg. de) and TargetScan TargetScanHuman 7.2 were utilized to predict miRNAs targeted for specific target genes. Additionally, the starBase database (http://starbase.sysu.edu.cn/index.php), miRNet, and miRcode database (http://www.mircode.org/) were used to predict the target long non-coding RNAs (lncRNAs) of miRNAs.

### 2.8 Mice

A total of 40 male C57BL/6 mice, each four weeks old, were acquired from Speford Biotechnology Company located in Beijing, China, and kept in animal facilities that were free from specific pathogens. All animal experiments adhered to the National Guidelines for the Ethics Review of Laboratory Animal Welfare. Ethical approval for all mouse experiments was granted by the Laboratory Animal Ethics Committee of the Beijing MDKN Biotech Company (MDKN-2023-054).

### 2.9 Virus isolation and purification

The bend3 cells were placed in an ultracentrifuge tube and subjected to centrifugation at 41,000 rpm for 2.5 hours at a temperature of 4 °C. After this step, the supernatant, devoid of cells, was removed and the sediment collected at the bottom was resuspended and rinsed with sterile phosphate-buffered saline (PBS). Sucrose solutions with concentrations of 10%, 25%, 45%, and 60% were prepared using PBS as the solvent. Specifically, three milliliters of 10% solution, three milliliters of 25% solution, two milliliters of 45% solution, and two milliliters of 60% solution were introduced into the centrifuge tubes. The virus solution was then carefully layered over the sucrose solutions. Following this, The samples underwent centrifugation at 27,000 rpm for 3 h at 4 °C with the aid of an SW41 rotor. Upon completion of centrifugation, three separate white bands appeared at the interfaces of the 10–25%, 25–45%, and 45–60% sucrose layers, which were subsequently aspirated and diluted with PBS. The samples were centrifuged once more at 27,000 rpm for 2 h using an SW41 rotor. After this round of centrifugation, the supernatant was removed and the resulting precipitate was resuspended in PBS before being stored in a refrigerator at −80 °C.

### 2.10 *In vivo* model construction

Mice were randomly divided into two groups: the mouse polyomavirus (MPyV) infection group and the control group. The MPyV infection group was constructed by injecting bend3 cells (1×10^6^/200μL), which express the MPyV intermediate antigen (mT), into the tail vein. The control group was administered 0.9% normal saline. One injection was administered on days one, four, and seven. One week after the final injection, mice were anesthetized with isoflurane and euthanized by cervical dislocation. Kidney specimens were collected for subsequent analysis.

### 2.11 Histological evaluation

The kidneys were preserved in 10% neutral buffered formalin and embedded in paraffin. Each paraffin section, measuring 3 μm in thickness, was stained with hematoxylin and eosin (H&E). An experienced renal pathologist examined the slides in a single-blind fashion to evaluate morphological signs of polyomavirus infection. Histopathological evidence of MPyV infection was characterized by the presence of viral inclusion bodies.

### 2.12 Reverse transcription qPCR (RT–qPCR)

Total RNA was extracted from the tissues using TRIzol reagent (Invitrogen, USA). RNA was reverse transcribed into complementary DNA (cDNA) using reverse transcriptase (Vazyme Biotech Co., Ltd.), followed by an RT–qPCR assay employing ChamQ Universal SYBR qPCR Master Mix (Vazyme Biotech Co., Ltd.). All quantified values were normalized based on the endogenous expression levels of GAPDH. Primer sequences used for RT–qPCR are listed in Table 1.

**Table 1.**
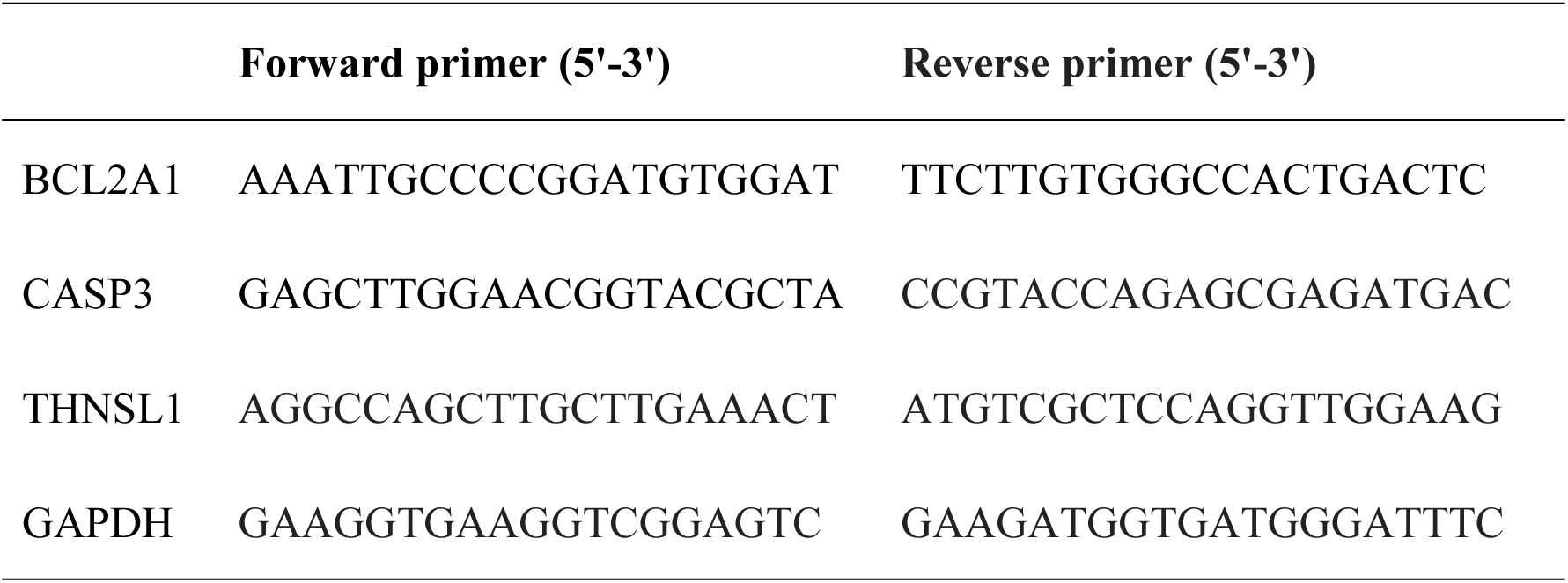
The sequences of the primers used for RT–qPCR.

### 2.13 Western blot

Kidney tissue was homogenized to extract total proteins using radioimmunoprecipitation assay (RIPA) lysis buffer (R0020; Solaibao). A BCA protein quantification kit (Beyotime, P0012) was used for protein quantification. The proteins were then separated by SDS-PAGE (Solaibao, P1200) and subsequently transferred onto a polyvinylidene fluoride (PVDF) membrane. Incubate The membrane with primary antibody at 4 °C overnight, followed by incubation with secondary antibody for 1 h. The primary antibodies used in this study were B-cell lymphoma 2-related protein A1 (BCL2A1) (Thermo, PA5-20268, 1:1,000), Caspase-3 (CASP3) (CST, 9662S, 1:1,000), threonine synthase like 1 (THNSL1) (SANTA, SC-100587, 1:1,000) and GAPDH (ZSGB-BIO, TA-08, 1:2,000). The secondary antibodies used were peroxidase-conjugated goat anti-rabbit IgG (ZSGB-BIO, ZB-2301, 1 : 5,000) or peroxidase-conjugated goat anti-mouse IgG (ZSGB-BIO, ZB-2305, 1 : 5,000). Visualization was performed using a chemiluminescence imager (Kechuang Ruixin Biotechnology Co., Ltd., K3000 mini). The optical density of the membrane was analyzed using the ImageJ software.

### 2.14 Statistical analysis

The Wilcoxon rank-sum test was used to compare differences in gene expression and immune cell infiltration between groups. Pearson’s correlation analysis was performed using the RP programming language. p<0.05 indicated that the difference was considered statistically significant. All statistical analyses were performed using R version 4.3.3.

## 3 Results

### 3.1 Identification of DEGs and enrichment analysis between BKPyVAN and STA samples

To identified DEGs between BKPyVAN and STA samples, we first merged GSE72925 and GSE47199 datasets as a training set after removing the batch effects via the “sva” packages in R packages (Figure 1A, 1B), and obtained a total of 13 BKPyVAN and 84 STA samples. There were 832 DEGs between BKPyVAN and STA samples in the training set, including 593 upregulated and 239 downregulated genes (Figure 1C). Heatmaps of the top 50 DEGs are shown in Fig. 1D. Concurrently, these DEG were used for GO and KEGG enrichment analyses. A total of 61 KEGG pathways were found to be significantly enriched (p < 0.05), with 766 significantly enriched BP entries, 59 significantly enriched MF entries, and 72 significantly enriched CC entries (Table S1). The top 15 significantly enriched KEGG pathways and the top five significantly enriched GO pathways are illustrated in Figure 1E and 1F.

**Figure 1.**
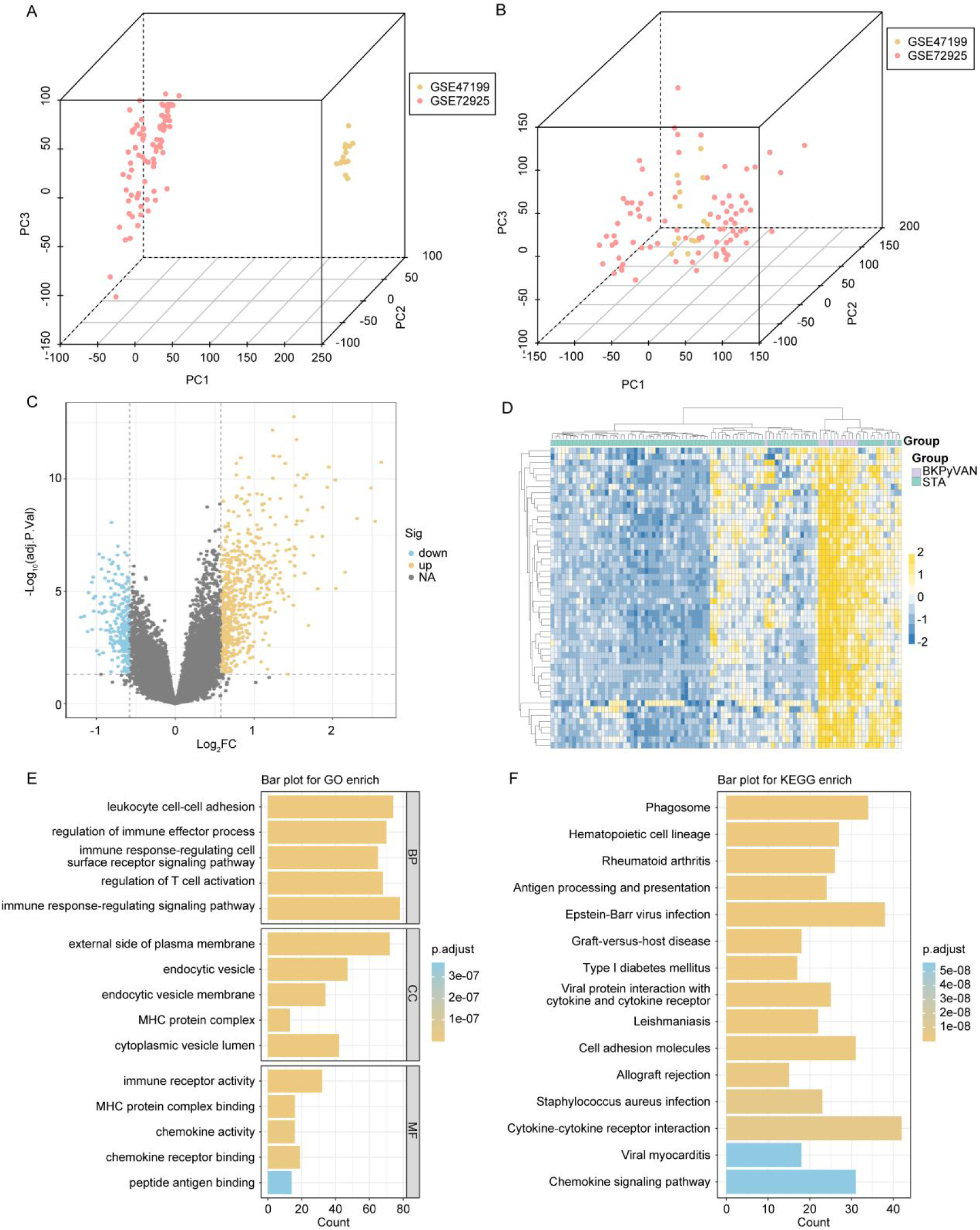
Identification of differentially expressed genes and enrichment analysis between BK polyomavirus-associated nephropathy (BKPyVAN) and stable grafts (STA) samples. Principal component analysis plot without batch effect elimination (A) and with batch effect elimination (B). C, The volcano plot of differentially expressed genes (DEGs) between BKPyVAN and STA samples in the training set. D, The heat map of top 50 DEGs between BKPyVAN and STA samples in the training set. The top 15 significantly enriched KEGG pathways (E) and the top 5 significantly enriched GO pathways (F).

### 3.2 Identification of BKPyV onset-related genes

To identify the genes closely related to BKPyVAN pathogenesis, we performed WGCNA using samples from the training set. We chose *β* = 12 as the optimal soft threshold for constructing scale-free networks (Figure 2A). WGCNA yielded 23 gene modules (Figure 2B). Among these 23 gene modules, two (magenta and yellow-green) were significantly related to BKPyVAN (Figure 2C, 2D). The correlations between module membership and gene importance are shown in Figure 2E and 2F. Finally, 672 genes found in magenta and yellow-green nodules were considered BKPyVAN pathogenesis-related genes for further study (Table S2).

**Figure 2.**
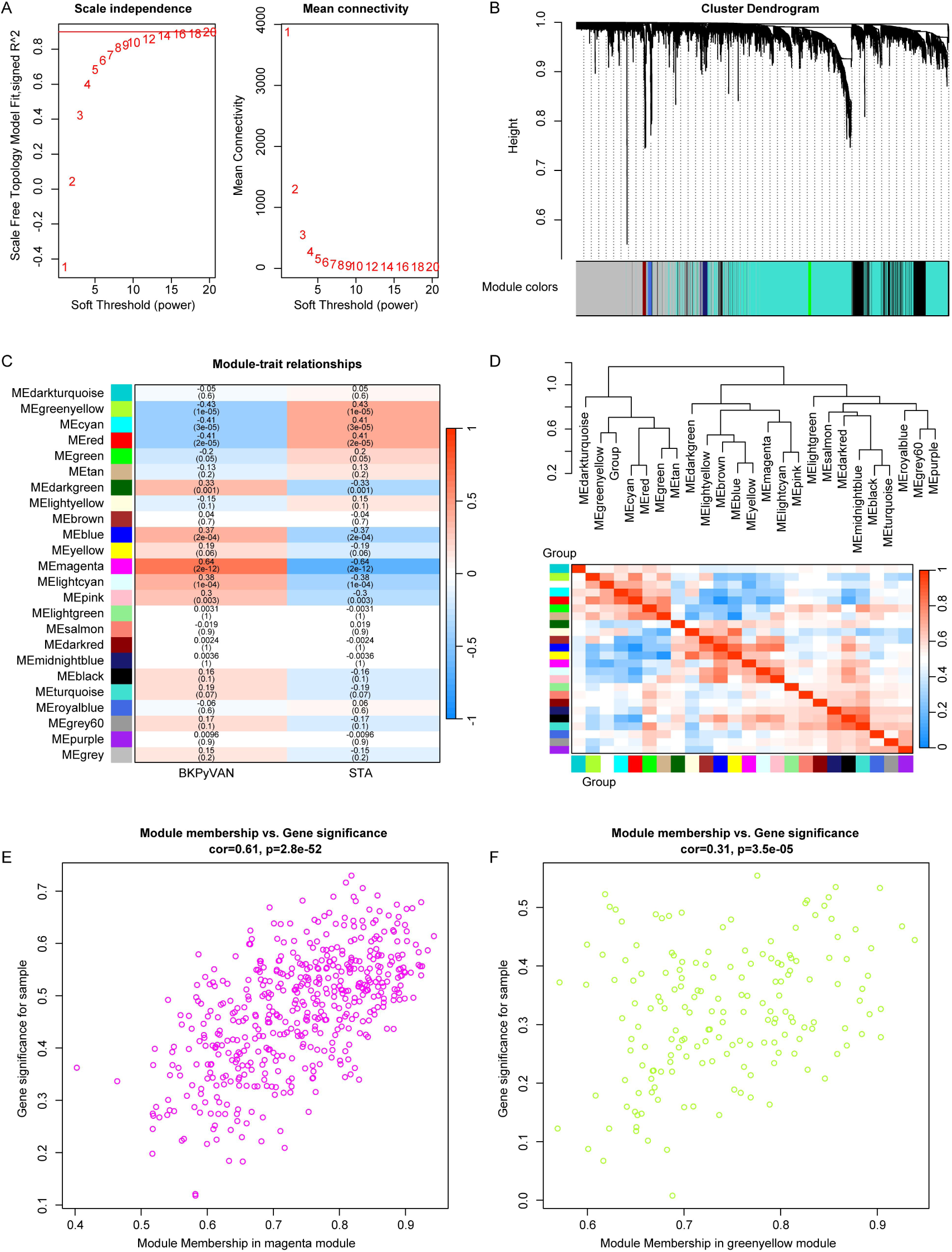
Identification of BKPyVAN onset-related genes. A, The optimal soft threshold *β*. B, The graph of clustering results of gene modules, the top half is the hierarchical clustering tree diagram of genes, the bottom half is the gene module. C, Heat map of correlation between modules and traits. D, Clustering tree and correlation heatmap between modules, the top figure shows the clustering tree of the modules after adding the traits, and the bottom figure shows the correlation heatmap between the traits and each module. Scatter plot showing the gene significance (GS) vs module membership (MM) for the magenta (E) and greenyellow (F) modules.

### 3.3 Identification of validation hub mitochondria-related genes in BKPyV

The genes in the modules screened by WGCNA were crossed with 832 DEGs and 1136 mitochondria-related genes, and nine genes were identified as target genes (Figure 3A, Table S2). The expression of the nine mitochondria-related genes in the BKPyVAN and STA groups is shown in Figure 3B.

**Figure 3.**
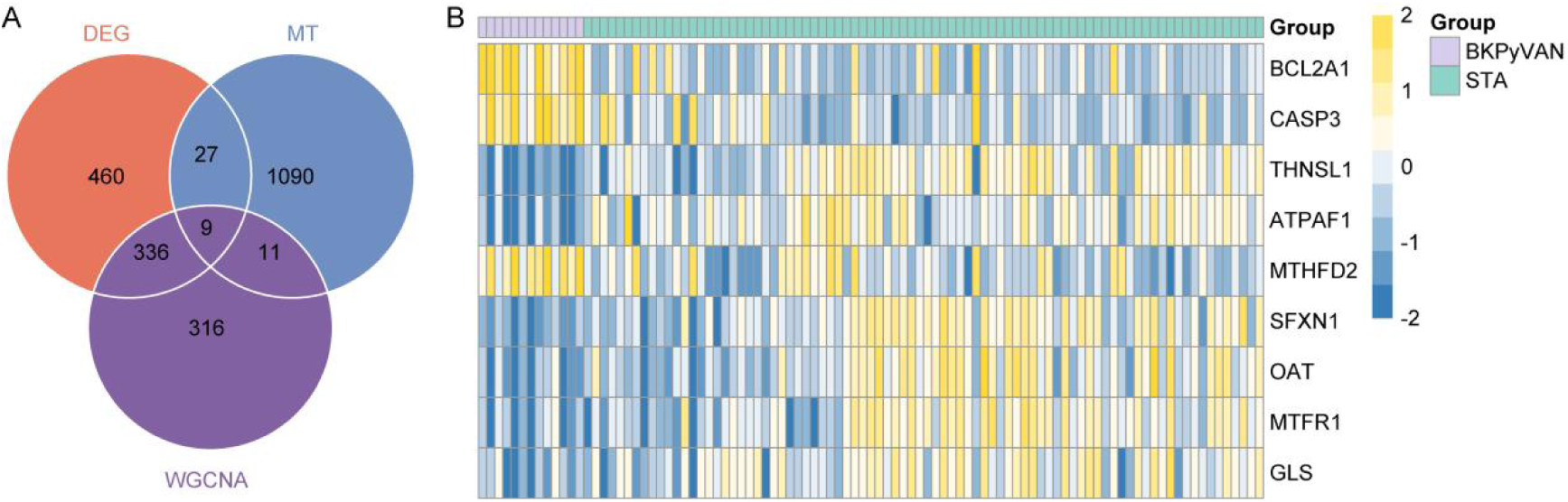
Identification of hub mitochondria-related genes in BKPyVAN. A, Overlapping genes among modules genes, DEGs, and mitochondria-related genes. B, Heat map of 9 mitochondria-related gene expression in the BKPyVAN and STA groups.

To identify robust mitochondria-related genes in BKPyVAN, we used multiple machine learning techniques to search mitochondria-related genes in BKPyVAN. Using RF, these nine genes were identified as potential BKPyVAN biomarkers (Figure 4A). Using lambdamine as a parameter, LASSO identified five features (*THNSL1*, *BCL2A1*, *CASP3*, *ATPAF1*, *SFXN1*) as potential biomarkers (Figures 4B, 4C). Three genes (*THNSL1*, *BCL2A1*, and *CASP3*) were identified using the RFE algorithm (Figure 4D). Finally, three overlapping genes, *THNSL1*, *BCL2A1*, *CASP3*, were identified and considered as robust biomarkers for BKPyVAN.

**Figure 4.**
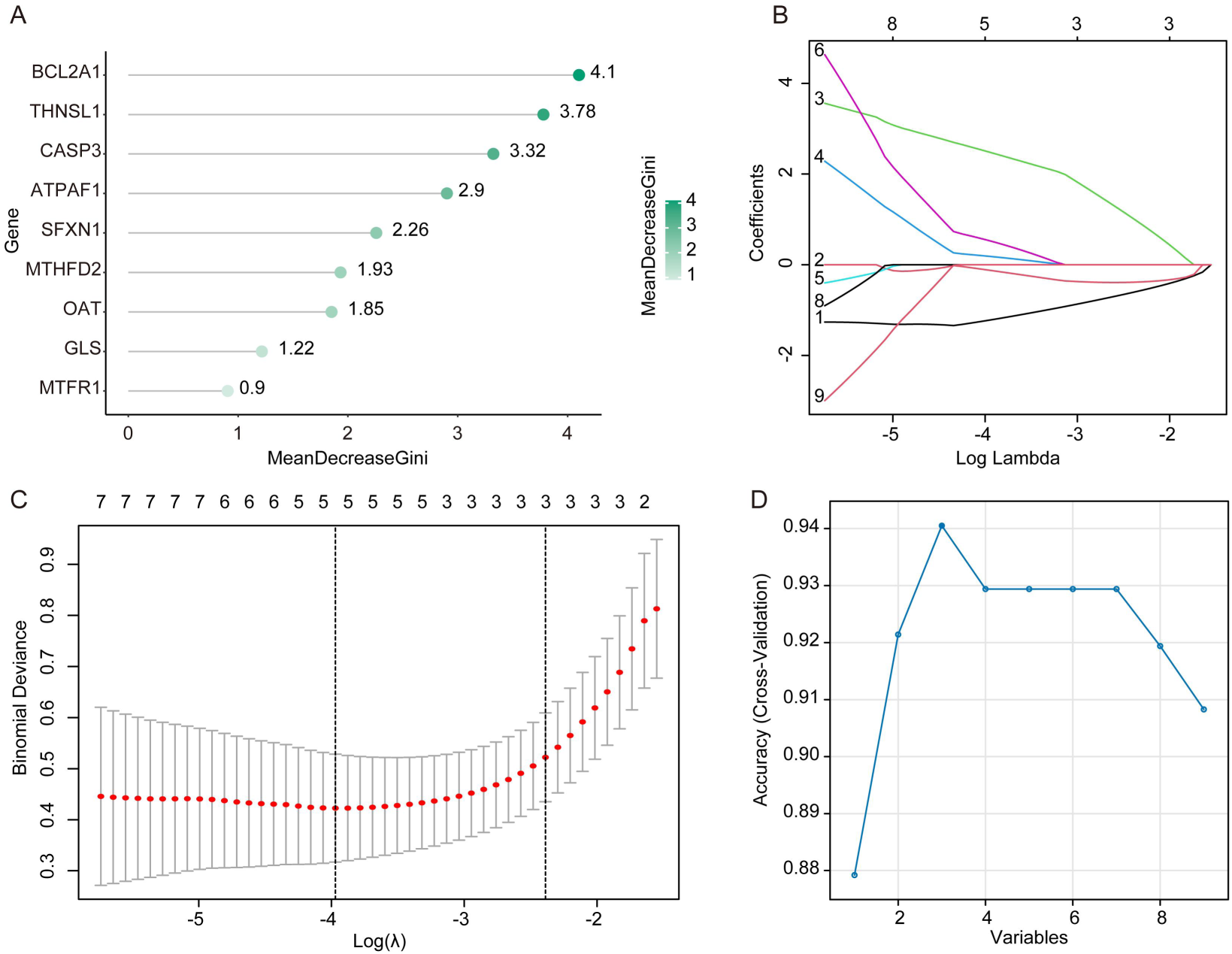
Identification of robust mitochondria-related genes in BKPyVAN. A, Random forest feature importance ranking. B, Regression coefficient path diagram, the x-axis represents the value of the regularization parameter (log), and the y-axis represents the size of the model parameters. C, Lasso regression cross-validation curve, the x-axis represents the value of the regularization parameter (log), and the y-axis is the likelihood deviation. D, Recursive feature elimination RFE, the abscissa represents the number of feature genes, and the ordinate represents the generalization error under 10-fold cross-validation.

### 3.4 The expression characteristics of *BCL2A1*, *CASP3*, and *THNSL1* in BKPyVAN

In the training set and two validation sets (GSE75693 and GSE120495), we analyzed the expression of *BCL2A1*, *CASP3*, and *THNSL1* between STA samples following kidney transplantation. As shown in Figure 5A, *BCL2A1* and *CASP3* were upregulated, whereas *THNSL1* was downregulated in BKPyVAN samples compared to STA in the training set. Consistent results were observed for the GSE75693 dataset (Figure 5B). In the GSE120495 dataset, *BCL2A1* was significantly upregulated in BKPyVAN samples, whereas *CASP3* and *THNSL1* exhibited trends of increased and decreased expression, respectively, in BKPyVAN samples. However, these findings were not statistically significant (Figure 5C). The lack of significance may be attributed to the small sample size.

**Figure 5.**
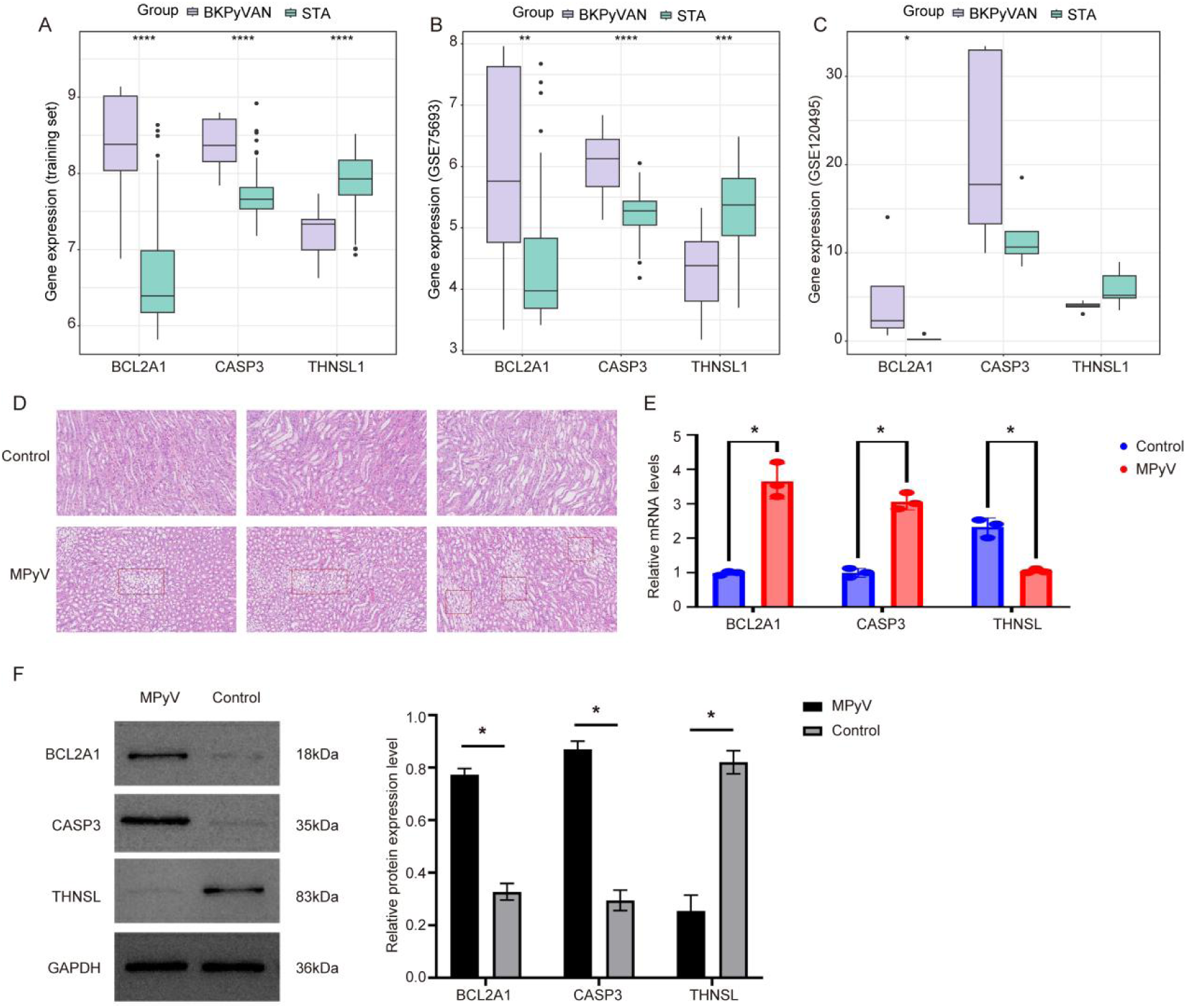
The expression characteristics of BCL2A1, CASP3, and THNSL1 in BKPyVAN. The expression of *BCL2A1*, *CASP3*, and *THNSL1* in BKPyVAN and STA samples in the training set (A), GSE75693 (B), and GSE120495 (C) datasets. D. Renal pathological manifestations in control and MPyV-infected mice. The expression levels of BCL2A1, CASP3, and THNSL1 mRNA (E) and protein (F) in the kidneys of virus-infected mice MPyV-infected mice. *p<0.05, **p<0.01, ****p<0.0001.

To further verify the expression levels of *BCL2A1*, *CASP3* and *THNSL1* in BKPyVAN, we established a mouse MPyV infection model via tail vein injection to simulate human BKPyV infection. Compared with the control group, mice infected with MPyV showed apparent kidney damage (Figure 5D). A portion of the renal parenchyma was damaged, resulting in blood plate formation. The walls of the renal tubules exhibited an abnormal shape, were reduced in thickness, and cell nuclei were not clearly discernible. Moreover, compared with the control group, the mRNA and protein expression levels of *BCL2A1* and *CASP3* were significantly increased in the kidneys of MPyV-infected mice, whereas the mRNA and protein expression levels of *THNSL1* were significantly decreased (Figures 5E and 5F). Uncropped original full-length western blot images are shown in Figure S1.

### 3.5 The diagnostic value and potential functional information of *BCL2A1*, *CASP3*, and *THNSL1* in BKPyVAN

To evaluate the diagnostic value of BCL2A1, CASP3, and THNSL1 in BKPyVAN, receiver operating characteristic (ROC) curves were plotted on the training set and two validation sets. As shown in Figure 6A, in the training dataset, the curve (AUC) values of *BCL2A1*, *CASP3*, and *THNSL1* were 0.929, 0.929, and 0.933, respectively. In the two validation sets, the AUC values for *BCL2A1*, *CASP3*, and *THNSL1* were greater than 0.75 (Figure 6B-6C). These results suggest that *BCL2A1*, *CASP3* and *THNSL1* may better differentiate BKPyVAN and STA patients.

**Figure 6.**
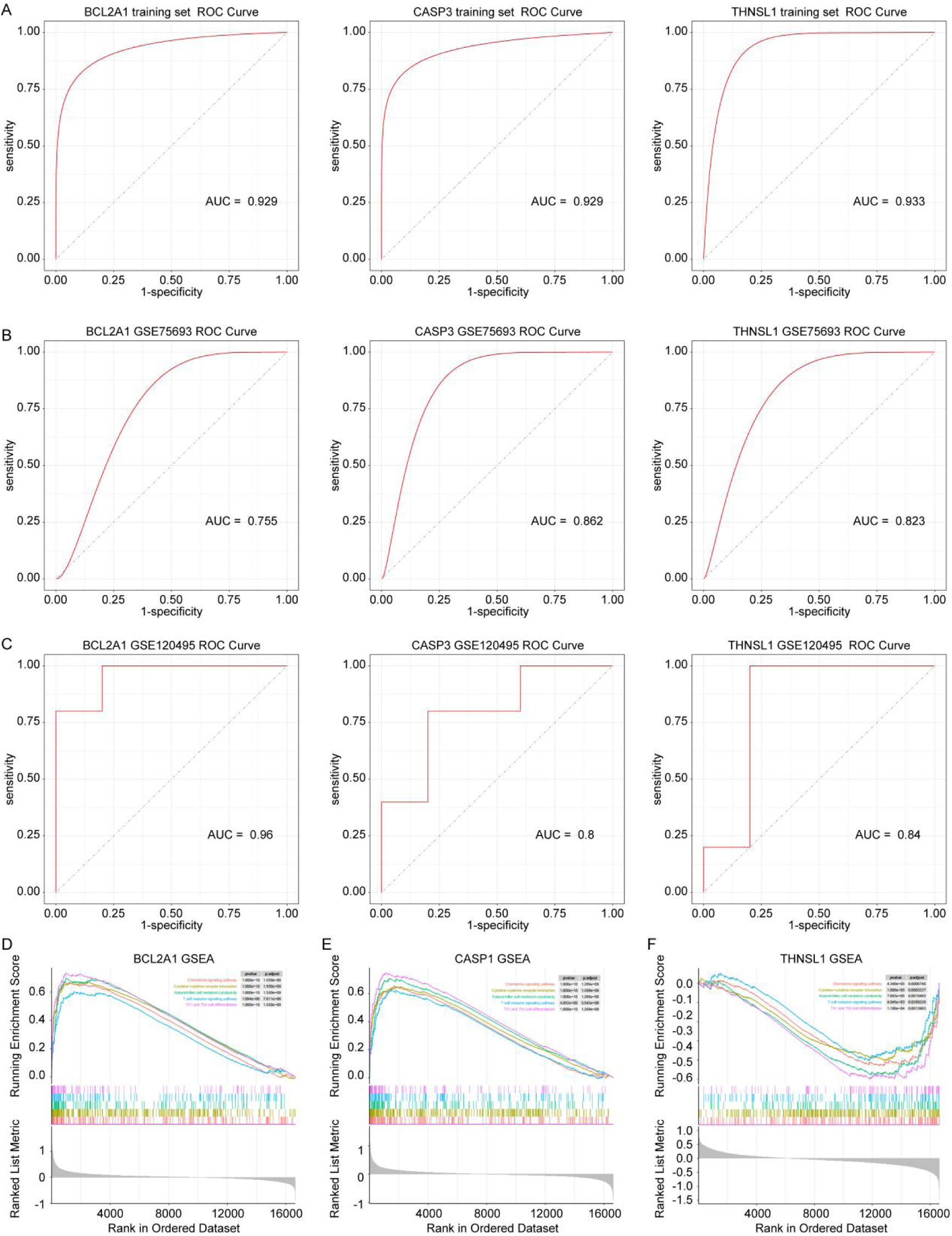
The diagnostic value and potential functional information of *BCL2A1*, *CASP3*, and *THNSL1* in BKPyVAN. The area under the curve (AUC) values of *BCL2A1*, *CASP3*, and *THNSL1* in the training set (A), GSE75693 (B), and GSE120495 (C) datasets. The potential functional information of *BCL2A1* (D), *CASP3* (E), and *THNSL1* (F) in BKPyVAN was performed using gene set enrichment analysis.

To explore the potential functional information of *BCL2A1*, *CASP3*, and *THNSL1* in BKPyVAN, we performed GSEA on the training set. We discovered that the chemokine signaling pathway, cytokine-cytokine receptor interaction, natural killer cell-mediated cytotoxicity, and T cell receptor signaling pathways were highly activated in BKPyVAN patients with high *BCL2A1* or *CASP3* expression (Figure 6D, 6E), while these signaling pathways were highly inhibited in BKPyVAN patients with low *THNSL1* expression (Figure 6F, Table S3). These results suggest that *BCL2A1*, *CASP3*, and *THNSL1* may be involved in BKPyVAN immune pathways.

### 3.6 *BCL2A1*, *CASP3*, and *THNSL1* was correlated with immune cell infiltration in BKPyVAN

To study the relationship between hub genes and immunity in BKPyVAN, we used the CIBERSORT algorithm to calculate the different immune cell infiltrates in the samples in the training set. As shown in Figure 7A, the proportion of macrophages M1, activated memory CD4+ T cells, T follicular helper (Tfh) cells, and gamma delta T cells was significantly increased in BKPyVAN samples compared to STA samples, whereas resting mast cells, activated natural killer (NK) cells, plasma cells, resting memory CD4+ T cells, naïve CD4+ T cells, and regulatory T cells (Tregs) were decreased. In the two validation sets, the relative content of Tfh cells was higher in the BKPyVAN samples than in the STA samples (Figure 7B, 7C). In addition, *BCL2A1* and *CASP3* expression was highly positively correlated with Tfh, while *THNSL1* expression was negatively associated with Tfh in the training set (Figure 7D-7F, p<0.05).

**Figure 7.**
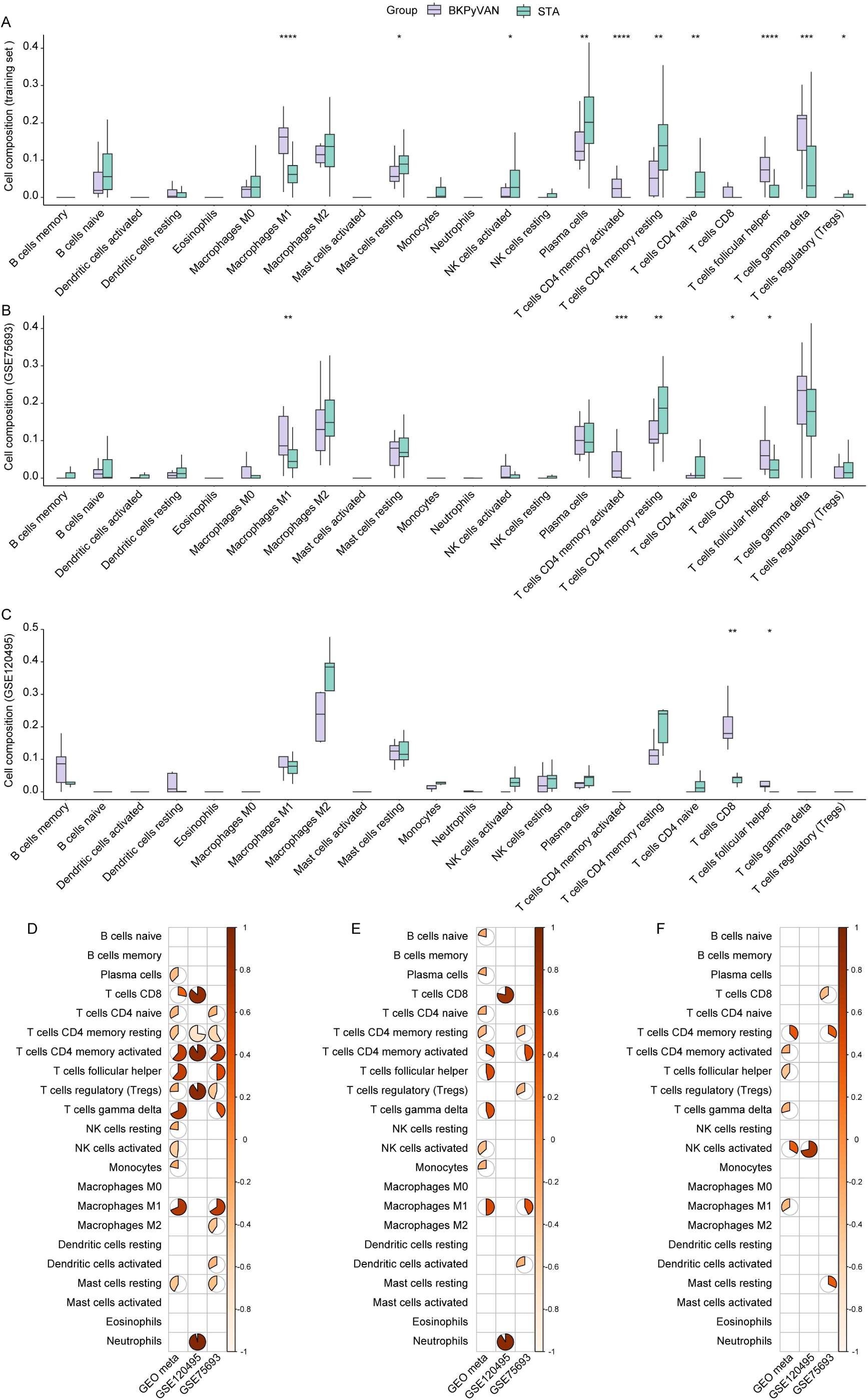
*BCL2A1*, *CASP3*, and *THNSL1* was correlated with immune cell infiltration in BKPyVAN. The proportion of 22 immune cell infiltration in BKPyVAN and STA samples in the training set (A), GSE75693 (B), and GSE120495 (C) datasets. The correlation between immune cell infiltration and *BCL2A1*, *CASP3*, and *THNSL1* expression in the training set (D), GSE75693 (E), and GSE120495 (F) datasets. *p<0.05, **p<0.01, ***p<0.01, ****p<0.0001.

### 3.7 Drug-target prediction

We conducted a drug target analysis focusing on *BCL2A1*, *CASP3*, and *THNSL1*. According to data from the GeneCards database (https://www.genecards.org/), OBATOCLAX MESYLATE is an inhibitor of *BCL2A1*. Furthermore, we found that 15 inferred drugs were associated with *BCL2A1*, 78 inferred drugs were linked to *CASP3*, and 2 drugs were related to *THNSL1*. The network figure illustrates these drug-target pairs (Figure 8A, Table S4).

**Figure 8.**
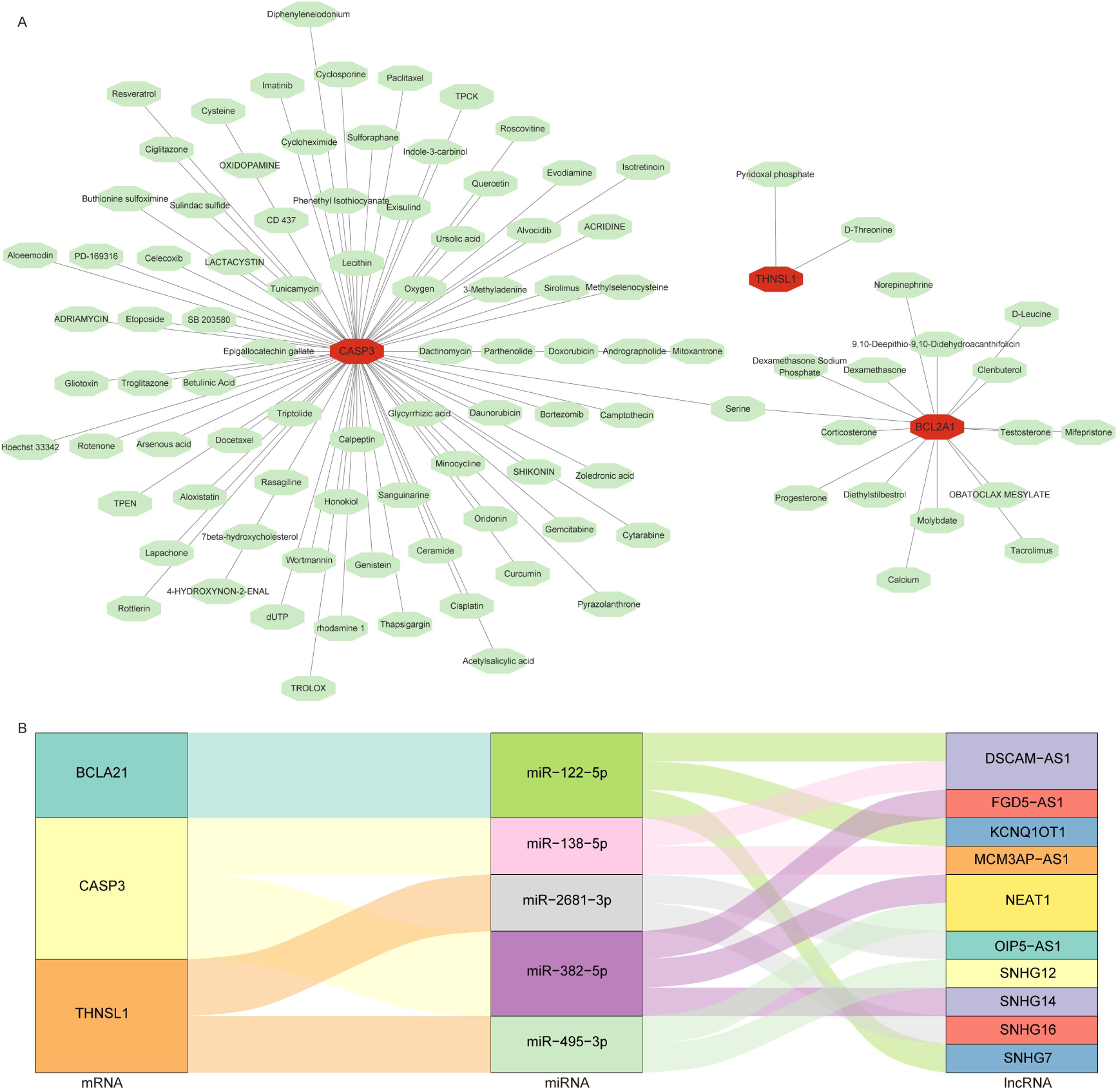
Prediction of hub genes-related drugs and construction of lncRNA-miRNA-mRNA regulating network. A, Drug and gene network diagram. B, lncRNA-miRNA-mRNA regulatory network.

### 3.8 Construction of lncRNA-miRNA-mRNA regulating network

To explore the regulatory mechanism of hub genes in BKPyV occurrence in renal transplantation, we first predicted the target miRNAs for *BCL2A1*, *CASP3*, and *THNSL1* using the miRDB, miRNet, and miRWalk databases. Five miRNAs (hsa-miR-122-5p, hsa-miR-138-5p, hsa-miR-382-5p, hsa-miR-495-3p, and hsa-miR-2681-3p) were obtained from the common prediction of the three databases. Subsequently, the Starbase database, miRNet, and miRcode database were utilized to predict potential long non-coding RNAs (lncRNAs) targeted by these five miRNAs By intersecting the lncRNAs identified across these three databases, a total of twelve lncRNAs were obtained. The selected miRNAs, lncRNAs, and three hub genes were plotted in a network using Cytoscape (Figure 8B; Table S5).

## 4 Discussion

Mitochondria are essential organelles within cells that perform multiple interrelated functions including the production of ATP and various biosynthetic intermediates. Additionally, they play a crucial role in cellular stress responses such as autophagy and apoptosis (12). Recent studies have revealed the critical role of mitochondrial dysfunction in BKPyVAN progression (6). Mitochondrial volume was reduced in the renal tubular cells of BKPyVAN patients (5), and mitochondrial fragmentation and p62/SQSTM1-mediated autophagy were also observed in allograft biopsies of BKPyVAN (7). However, the diagnostic value of mitochondria-related genes for BKPyVAN remains unclear. In the current study, we identified three mitochondria-related biomarkers (*THNSL1*, *BCL2A1*, and *CASP3*) in BKPyVAN by integrating bioinformatics and machine learning techniques, which may serve as potential targets for the diagnosis and treatment of BKPyVAN. These genes showed significant diagnostic value and were correlated with immune cell infiltration, suggesting their critical role in the pathogenesis of BKPyVAN.

The L-threonine synthase gene, threonine synthase-like 1 (*THNSL1*), is a paralog of threonine synthase like 2 (*THNSL2*). THNSL1 is expressed in oral squamous cell carcinoma (OSCC) cells (13) and may be correlated with phantom tooth pain (14). THNSL2 expression in de novo secreted osteoclast factor (SOFAT)-activated T cells plays an important role in bone remodeling and mediates bone resorption in diseases, such as rheumatoid arthritis and periodontitis (15). However, the role of *THNSL1* and *THNSL2* in kidney disease has not been reported. In this study, we found that *THNSL1* was upregulated in BKPyVAN and its expression could differentiate BKPyVAN and STA patients, suggesting that *THNSL1* might serve as a potential biomarker for the diagnosis and monitoring of BKPyVAN.

B-cell lymphoma 2-related protein A1 (BCL2A1) belongs to the BCL-2 family and consists of essential regulators that primarily oversee the release of cytochrome c from mitochondria during the intrinsic apoptotic pathway (16). As a vital regulator of the mitochondria-mediated apoptotic pathway, BCL2A1 plays a key role in this process (17). In the mitochondria, endogenous TRIM28 interacts with BCL2A1, and reducing TRIM28 levels leads to decreased ubiquitination of BCL2A1 (18). Previous research has demonstrated that BCL2A1 is significantly linked to tumorigenesis, patient prognosis, and chemotherapy resistance in various tumors (19–21). Furthermore, *BCL2A1* was found to be upregulated in diabetic nephropathy and correlated with the infiltration of immune cells (22). This finding was consistent with our research results, which indicated that *BCL2A1* is highly expressed in BKPyVAN and is associated with the infiltration of T cells. BCL2A1 has been identified as a target gene influenced by nuclear factor kappa B (NF-κB) (16), and the NF-κB signaling pathway is the primary mechanism that facilitates replication of mouse polyomavirus (23). Moreover, nuclear factor of activated T cells (NFAT) promotes BKPyV infection, and NFAT regulates BKPyV transcription by cooperating with the transcription factors AP-1 and NF-κB (24). Interestingly, the NF-κB signaling pathway was activated in BKPyVAN patients with high *BCL2A1* expression levels. Accordingly, we hypothesized that in BKPyVAN patients, *BCL2A1* might affect T cell function by regulating the NF-κB signaling pathway, thereby affecting BKPyV infection or transcription.

Caspase-3 (*CASP3*) is a critical cysteine protease that plays a central role in apoptosis (25). As an effector enzyme, it is activated in various cell death signaling pathways and facilitates programmed cell death (26). Research has indicated that the activity of CASP3 was closely associated with the onset and progression of several kidney diseases, where apoptosis constitutes a significant pathological process. Studies have shown that inhibiting CASP3 activity can reduce tubular apoptosis and slow the progression of kidney disease (27). For example, in models of polycystic kidney disease (PKD), inhibition of caspase-3 decreased tubular cell apoptosis and proliferation, thereby improving renal function (28). Furthermore, excessive activation of CASP3 exacerbates renal tubular damage and accelerates fibrosis progression, ultimately leading to a decline in renal function (29, 30). In diabetic mice, *CASP3* inhibition results in improved renal function and reduced urinary albumin excretion (27). In the present study, we observed that *CASP3* expression was decreased in BKPyVAN and closely correlated with immune cell infiltration. *CASP3* plays a multifaceted role in kidney diseases. Regulation of *CASP3* activity may offer novel strategies for the treatment of these conditions. Future research should investigate the specific mechanisms of *CASP3* across various types of kidney diseases (such as BKPyVAN) to facilitate the development of more effective therapeutic approaches.

This study identified mitochondrial genes (*THNSL1*, *BCL2A1*, and *CASP3*) associated with BKPyVAN, and explored their diagnostic value and immunoregulatory roles. However, this study has several limitations must be acknowledged. First, the samples utilized in this study were primarily sourced from public databases, resulting in limited data volume that may compromise the statistical power and generalizability of the findings, thereby necessitating validation with larger cohorts. Second, while the expression changes of *THNSL1*, *BCL2A1*, and *CASP3* correlated with BKPyVAN, their specific molecular mechanisms in viral nephropathy remain unvalidated through in vitro or in vivo experimental models. Finally, the correlation of *THNSL1*, *BCL2A1*, and *CASP3* with immune cell infiltration was inferred through bioinformatic analyses, lacking direct experimental confirmation, such as flow cytometry or immunohistochemistry, to substantiate specific alterations in immune cell populations.

## 5 Conclusion

This study identified three robust mitochondria-related biomarkers, *THNSL1*, *BCL2A1*, and *CASP3*, which were associated with BKPyVAN following kidney transplantation. These biomarkers were differentially expressed in BKPyVAN samples, with *THNSL1* and *BCL2A1* being upregulated and *CASP3* being downregulated. They demonstrated significant diagnostic value, as indicated by the ROC curve analysis, and were implicated in immune pathways. Furthermore, a correlation between these genes and immune cell infiltration in BKPyVAN has been established. These findings suggest that *THNSL1*, *BCL2A1*, and *CASP3* may play critical roles in BKPyVAN pathogenesis and may serve as potential targets for diagnostic and therapeutic interventions.

## Declarations

## Acknowledgement

Not applicable.

## Ethical approval and consent to participate

All animal experiments adhered to the National Guidelines for the Ethics Review of Laboratory Animal Welfare. Ethical approval for all mouse experiments was granted by the Laboratory Animal Ethics Committee of the Beijing MDKN Biotech Company (MDKN-2023-054).

## Consent for publication

Not applicable.

## Data Availability Statement

Data supporting the conclusions of this study are openly available in the Gene Expression Omnibus (GEO, https://www.ncbi.nlm.nih.gov/geo/) database, with the following IDs: GSE72925, GSE47199, GSE120495, and GSE75693.

## Authors’ contributions

Changhai Xu participated in the design of this study, and Qin Feng performed statistical analysis. Xueying Wang, Haibo Wu, Wei Li, Fei Lin, Na Lin, Shiyin Shen, Shubin Pan, Tong Chen, Donghui Zhang and Long He carried out the study and collected background information. Yan Cui drafted the manuscript. All authors have read and approved the final manuscript.

